# An Explainable Deep Learning Framework for Imaging Genetics: Deriving Brain-Genotype Scores to Link Genetic Variation, Brain Structure, and Cognition

**DOI:** 10.64898/2026.05.06.26352595

**Authors:** Kholod Thaker Alhasani, Upamanyu Ghose, Joshua Sammet, Taiyu Zhu, Sihao Xiao, Benoit Hastoy, Paul Brennan, Karen Froud, Brittany Ulm, Cornelia van Duijn, Laura M Winchester, Brian D. Marsden, Alejo Nevado-Holgado

## Abstract

Imaging genetics aims to understand how genetic variation influences brain structure and cognitive function. Traditional approaches often rely on imaging-derived phenotypes (IDPs), which reduce high-dimensional brain images to predefined summary measures and may miss subtle or spatially distributed genotype-related effects. We developed brain-genotype scores, continuous image-based representations of genetic variation learned directly from structural MRI. Using a multitask deep-learning framework trained on T1-weighted MRI from the UK Biobank, we predicted genotype dosage for 120 SNPs. The resulting genotype probability estimates were used as continuous brain-genotype scores. Unlike conventional IDPs, these scores are learned directly from raw images and capture distributed neuroanatomical patterns associated with genetic variants. Gradient-based saliency maps were used to localise neuroanatomical regions contributing to each score, providing interpretable links between genetic variation and brain anatomy. To evaluate their biological relevance, brain-genotype scores generated from an independent, held-out test set were used as neuroanatomical markers in association analyses with seven cognitive phenotypes, adjusted for population structure and technical covariates. A total of 147 out of 840 score-based associations survived FDR correction. In contrast, corresponding analyses using the original genotype data and traditional machine-learning-based scores trained on IDPs identified only two significant associations in total. These findings suggest that brain-genotype scores capture cognition-related neuroanatomical information beyond that available from genotype data alone, providing an interpretable, individual-level representation of genotype-related brain variation. By encoding both genetic and neuroanatomical information in SNP-specific brain-genotype scores, the framework may extend beyond cognition to other phenotypes, offering a new route to genotype-phenotype discovery in imaging genetics.

**Key Messages:**

- Brain-genotype scores are MRI-derived representations of genetic variation learned directly from structural brain images.
- The scores integrate genetic and neuroanatomical information into SNP-specific biomarkers for downstream phenotype analyses.
- Saliency-map analyses provide interpretable, SNP-specific neuroanatomical signatures underlying brain-genotype scores.
- Brain-genotype scores showed substantially more significant associations with cognitive phenotypes than genotype data.
- This framework provides a new approach for discovering genotype–phenotype relationships in imaging genetics.

## Introduction

Imaging genetics seeks to elucidate the complex relationships between genetic variation, brain structure, and cognitive function. By integrating neuroimaging and genomic data, it enables the identification of biomarkers relevant to neurodegenerative diseases, ageing, and cognitive traits [5, 4], while providing insights into the biological pathways underlying brain function and pathology [22].

Most large-scale imaging genetics studies use genome-wide association study (GWAS) frameworks to identify genetic variants associated with predefined imaging-derived phenotypes (IDPs) [10, 35, 41]. While these studies have substantially advanced our understanding, they face several methodological challenges that constrain their ability to capture the full complexity of brain-gene relationships. GWAS analyses have traditionally relied on statistical models that assume additive and linear relationships between single–nucleotide polymorphisms (SNPs) and brain phenotypes [39, 44]. Such assumptions may overlook non-linear effects or higher-order interactions across brain regions [29], although recent advances, including whole-genome regression and machine-learning-based methods, have substantially improved scalability and statistical efficiency for large biobank datasets [25]. When applied to imaging genetics, these methods still require complex neuroimaging data to be summarised into low-dimensional phenotypes to meet statistical and computational constraints. As a result, high-dimensional MRI data are commonly reduced to summary IDPs, often using dimensionality-reduction techniques such as principal component analysis (PCA) or independent component analysis (ICA) [10, 48]. While practical, this process risks discarding biologically meaningful spatial patterns not captured by predefined feature sets or regional metrics [44, 29]. Finally, many studies adopt a univariate analysis strategy, limiting the detection of shared or distributed genetic influences across brain traits and potentially obscuring the polygenic and pleiotropic nature of brain-genetic relationships, particularly when genetic variants exert small, coordinated effects across different brain regions [43].

To address these limitations, alternative approaches are emerging. One promising direction is reverse imaging genetics, in which models predict genetic variation from brain imaging data. These approaches can enhance sensitivity for detecting genotype-phenotype associations, especially when incorporating multiple imaging traits or multimodal data [30, 13, 45, 24]. Building on this direction, we introduce brain-genotype scores, image-based representations of genetic variation learned directly from structural MRI. Unlike conventional approaches that rely on predefined IDPs, brain-genotype scores capture distributed genotype-related neuroanatomical patterns directly from high-dimensional brain images without handcrafted regional features or dimensionality reduction. To derive these scores, we employ an explainable multi-task convolutional neural network (CNN) framework capable of leveraging the spatial complexity of 3D brain images and modelling both linear and non-linear gene–brain relationships [20, 3, 17]. We evaluate brain-genotype scores as quantitative neuroanatomical markers in association analyses with cognitive phenotypes, including fluid intelligence, memory, and reaction time. We further compare their utility with analyses based on the original SNP genotypes and conventional IDP-based approaches. Finally, we investigate the neuroanatomical patterns underlying the scores through model interpretability analyses. By integrating imaging and genetic information into SNP-specific image-based representations, the proposed framework provides a data-driven approach for identifying biologically meaningful genotype–phenotype associations and uncovering complex relationships between genetic variation, brain structure, and cognition.

## Methods

### Datasets and preprocessing

We used T1-weighted 3D brain MRI scans from the UK Biobank (UKB) dataset (UKB field ID 20252 as of February 2023 under UKB application 15181). All images had undergone the standard UKB preprocessing pipeline, including skull stripping, brain extraction, and non-linear registration to MNI152 standard space, as described in [2]. No additional preprocessing was performed. To minimise population stratification effects, analyses were restricted to subjects of White British ancestry, as defined by UKB genetic ancestry field ID 22006. All White British subjects with available T1-MNI152 space MRI images were included, yielding a total of 41,382 subjects. Demographic characteristics of the full dataset are summarised in Table 1. For the CNN model, the dataset was divided into training, validation, and test subsets (80%, 10%, and 10%) as summarised in Table 2. The testing subset was exclusively used for downstream statistical analyses and not for making choices on CNN architecture or algorithmic design. Additionally, IDPs and demographic variables (including age and sex), and genetic principal components (PCs) for the same subjects, available in the UKB, were used for downstream statistical and machine-learning analyses described below. For genetic data, we selected SNPs previously associated with brain structural variation. For each SNP, genotype served as the target label for CNN training. These SNPs were encoded into three genotype classes: 0 (*bb*, two minor alleles), 1 (*Ab*, heterozygous), and 2 (*AA*, two major alleles). To identify relevant SNPs, we selected variants reported in large-scale GWAS as significantly associated with T1-weighted IDPs [10, 41]. Linkage disequilibrium pruning (r=0.2) and a minor allele frequency threshold of 0.05 were applied, and SNPs lacking representation in any genotype class within the training, validation, or test subsets were excluded. After filtering, 120 SNPs were retained.

**Table 1.** Demographic Data for Full Dataset.

| Age (years) |  |  |
| --- | --- | --- |
| mean | 64.9 |  |
| std | 7.6 |  |
| min | 46 |  |
| 25% | 59 |  |
| 50% | 65 |  |
| 75% | 71 |  |
| max | 85 |  |
| Sex | Count | Percentage (%) |
| Female | 21 467 | 51.88 |
| Male | 19 912 | 48.12 |
| Assessment centre | Count | Percentage (%) |
| 11025 | 20 476 | 54.42 |
| 11027 | 9 961 | 26.48 |
| 11026 | 5 942 | 15.79 |
| 11028 | 1 244 | 3.31 |

**Table 2.**
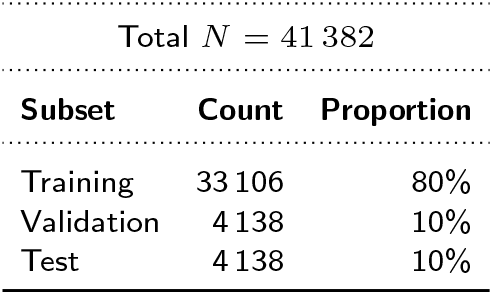
Summary of the CNN Model Dataset Split.

### CNN model and Derivation of Brain-Genotype Scores

A lightweight 3D convolutional neural network (CNN) was developed to generate brain-genotype scores from T1-weighted MRI scans (Figure 1). The model architecture used a modified ResNet-10 backbone[15], adapted for volumetric data by replacing standard 2D operations with their 3D equivalents. The model input consisted of MRI volumes with dimensions of 182 × 218 × 182 voxels and 1 mm isotropic resolution.

**Figure 1.**
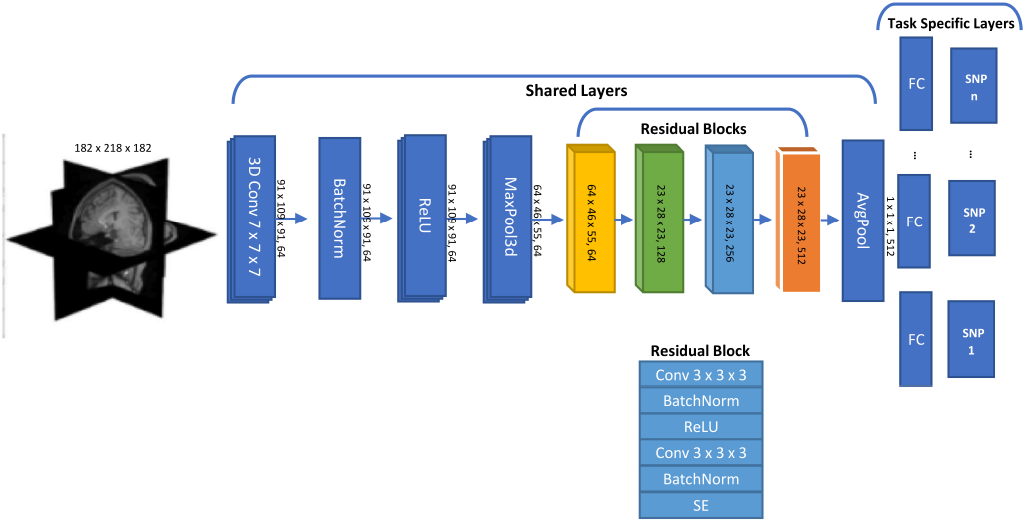
3D ResNet-10 model architecture

The learning task was formulated as a multi-task, probabilistic prediction problem, where the CNN simultaneously generated genotype probability distributions for multiple SNPs for each subject. Each SNP represented an independent output task with three possible genotype classes: 0 (two copies of the minor allele), 1 (heterozygous), or 2 (two copies of the major allele). This multi-task formulation allowed the network to learn shared feature representations across SNPs while producing SNP-specific probabilistic outputs (brain-genotype scores), which sped up learning and final prediction per SNP. To balance computational feasibility and learning stability, we explored different task group size (1, 10, 100, and 1,000 SNPs per model) during model development using randomly sampled SNPs. Larger task sets increased computational cost and slowed convergence. Based on this trade-off, we selected a configuration predicting 10 SNPs simultaneously. Consequently, the network produced an output tensor of shape *T ×* 3, where *T* = 10 is the number of SNPs, with each row containing predicted class probabilities for each SNP genotype. An overview of the model architecture is illustrated in Figure 1.

The CNN architecture began with an initial 7×7×7 convolutional layer, followed by batch normalisation[18], ReLU activation and max pooling[20]. Subsequently, four residual blocks were included, each containing two 3×3×3 convolutional layers with batch normalisation and ReLU activation after each convolution. To enhance feature representation, Squeeze-and-Excitation (SE) modules[16] were integrated into each residual block. These SE modules applied channel-wise attention, enabling the model to recalibrate feature responses and focus on informative channels. Each SE block first applied global average pooling to generate a compact descriptor of each channel. This descriptor was passed through two fully connected layers with a reduction ratio of 16 and non-linear activations to compute a set of channel-wise weights, which were then used to rescale the original feature maps. Mathematically, each SE block is defined as follows:

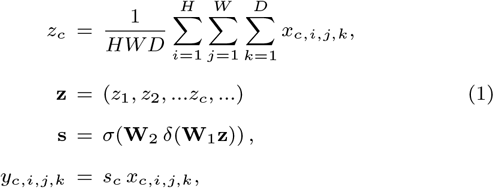

where *z*_*c*_ (scalar) is the average voxel activation for channel *c*; *H, W, D* (scalars) are the height, width, and depth of the feature map; *x*_*c,i,j,k*_ (scalar) is the activation at spatial location (*i, j, k*) in channel *c* of the input feature map; **z** (1D vector) is the vector of channel-wise pooled activations; **s** is the vector with the weights given to each channel *c*; **W**_1_ and **W**_2_ are the weights of the two fully connected layers, where *δ*(*·*) and *σ*(*·*) denote the ReLU and sigmoid activation functions, respectively; *y*_*c,i,j,k*_ (scalar) represents the output activation at spatial location (*i, j, k*) in channel *c* of the resulting feature map; and *s*_*c*_ (scalar) the weight given to channel *c*. After feature extraction, we applied global average pooling, resulting in a 512-dimensional vector per subject. This vector was passed to task-specific fully connected heads, each outputting the softmax probabilities for one SNP genotype—referred to as the brain-genotype score. These probability-based scores were later used as quantitative neuroanatomical markers in downstream association analyses with cognitive outcomes. As the three class probabilities sum to one, one probability was omitted to avoid perfect multicollinearity, and the remaining two were used as predictors in the downstream regression analyses. To ensure their reliability as probabilistic biomarkers, calibration was assessed using expected calibration error and reliability diagrams (Supplementary Figure 4). To assess the relationship between CNN-derived brain-genotype scores and the corresponding actual genotypes, the three genotype probabilities were summarised as expected genotype dosage, calculated as 0 *× p*_0_ + 1 *× p*_1_ + 2 *× p*_2_, where *p*_0_, *p*_1_, and *p*_2_ represent the predicted probabilities of genotype classes 0, 1, and 2, respectively. Pearson correlation was then used to quantify the relationship between expected and actual genotype dosage across the held-out test data. The CNN was implemented in *Python v3.10.0* using *PyTorch v1.13.1*, and trained using the AdamW optimiser with cross-entropy loss computed individually for each SNP output. Per-task losses were summed to form a single multi-task objective for model optimisation. Hyperparameter tuning was performed using Optuna (50 trials) to optimise learning rate, batch size, and weight decay. The best configuration used a learning rate of 5 *×* 10^*−*6^, batch size of 20, and weight decay 8.01*×*10^*−*5^, with equal weighting applied across all tasks to maintain an assumption-free multi-task learning setup. Finally, we trained 12 separate models, each predicting a unique subset of 10 SNPs, collectively covering all 120 selected variants as detailed in the “Datasets and Preprocessing” Section. All CNN models were trained on two NVIDIA GeForce RTX 3090 GPUs using CUDA 12.8 with data-parallel training.

#### Alt text

Diagram of the 3D ResNet-10 convolutional neural network used to generate brain-genotype scores from T1-weighted MRI scans. The network consists of an input MRI volume, an initial convolutional layer, batch normalisation, ReLU activation, max pooling, and four residual blocks with squeeze-and-excitation modules. Features are aggregated using global average pooling and passed to task-specific fully connected output layers that predict genotype probabilities for multiple SNPs simultaneously.

### Sex prediction task

To evaluate the generalisability of the CNN architecture to other biological prediction tasks, we assessed its ability to predict sex from T1-weighted brain MRI. Sex prediction is commonly used as a benchmarking task in neuroimaging [38], as it explains a substantial portion of variance in brain structure [26]. We used the same MRI input data, preprocessing pipeline, and training parameters as in the brain-genotype modelling task. The sex prediction task was incorporated into the multi-task learning framework and trained jointly with nine SNP modelling tasks, allowing evaluation of the architecture’s capacity to handle multiple biological prediction tasks simultaneously.

### Association Between Brain-Genotype Scores and Cognitive Performance

To assess whether CNN-derived brain-genotype scores capture variance relevant to cognition, we tested their associations with individual differences in cognitive performance using linear regression models. Cognitive phenotypes were obtained from the UK Biobank and were restricted to measures collected at the baseline assessment visit. All cognitive tests were administered in person using a touchscreen interface following UK Biobank protocols, except for online numeric memory, which was administered online via a web-based platform. Cognitive measures and their corresponding UK Biobank field IDs included fluid intelligence (20016), reaction time (20023), numeric memory (4282), numeric memory online (20240), matrix pattern puzzles (6373), symbol digit matches (23324), and tower rearranging (21004). Reaction time values were log-transformed prior to analysis to reduce positive skewness. All other cognitive measures were analysed in their original scale. As confounding factors, we included age, sex, education scores, and the top ten genetic principal components (PCs), which have become standard in the field [42, 7, 8]. Only participants with both imaging and cognitive data available were included in the association analyses, therefore, effective sample size varied across models. For each cognitive test, we used a nested model comparison framework: (i) a full model including the two retained probability components and confounders, and (ii) a reduced model with confounders only. An F-test based on the residual sum of squares (RSS) comparison between the two models tested the joint contribution of the probability components, indicating whether inclusion of the brain-genotype score significantly improved model fit. The resulting p-value therefore reflects the overall association between each brain-genotype score and the cognitive phenotype, rather than the contribution of either probability component individually.

To quantify effect size, we calculated the change in *R*^2^ (Δ*R*^2^) between the full and reduced models. This change represents the additional variance in cognitive performance that is explained by brain-genotype scores beyond what can be accounted for by confounders. This provided a unified measure of the explanatory utility of the brain-genotype scores across cognitive domains. Datasets used in this analysis are summarised in Table 3. We further explored the influence of global head-size on the observed associations by repeating the regression models with head-size scaling included as an additional covariate. This analysis evaluated how adjusting for overall brain volume affected the magnitude and significance of brain-genotype–cognition associations.

**Table 3.** Demographic data for the linear regression models dataset used in cognitive outcome analyses. This dataset corresponds to the test set of the CNN models used to generate brain-genotype scores.

| Total $N = 4139$ | | |
| --- | --- | --- |
| Age (years) |  |  |
| mean | 64.8 |  |
| std | 7.6 |  |
| min | 46 |  |
| 25% | 59 |  |
| 50% | 65 |  |
| 75% | 71 |  |
| max | 83 |  |
| Sex | Count | Percentage (%) |
| Female | 2152 | 51.99 |
| Male | 1987 | 48.01 |
| Assessment Centre | Count | Percentage (%) |
| 11025 | 2012 | 53.8 |
| 11027 | 999 | 26.7 |
| 11026 | 606 | 16.2 |
| 11028 | 123 | 3.3 |

### Reproducibility of brain-genotype scores

CNN training is stochastic, and model predictions may vary across random initialisations. To assess the stability of CNN-derived brain-genotype scores, we computed pairwise correlations between outputs from models trained with different random seeds. For each SNP, Pearson correlation coefficients were calculated using aggregated genotype probability vectors across subjects. To evaluate reproducibility as a function of model confidence, subjects were filtered using confidence thresholds ranging from 0.1 to 0.9, and correlations were recomputed at each threshold. Correlation values were visualised as heatmaps, together with the corresponding number of retained subjects. SNPs showing significant associations with cognitive phenotypes (FDR-corrected p-value) were highlighted to assess whether cognitively relevant brain-genotype scores exhibited greater stability. To summarise reproducibility, mean correlations across thresholds were computed for each SNP and compared between significant and non-significant SNPs using a Mann–Whitney U test. Robustness was further assessed using permutation testing (2,000 iterations) and bootstrapped 95% confidence intervals (2,000 resamples). Group differences were visualised using box plots of mean correlation values.

### Comparative Analyses with Classical ML Models and Actual Genotypes

To benchmark the CNN-derived brain-genotype scores, we conducted comparative analyses using (i) genotype probability scores generated by classical machine learning (ML) models trained on imaging-derived phenotypes (IDPs), and (ii) the actual genotype dosage. In both cases, the resulting scores or genotype values were tested for associations with cognitive performance using the same statistical framework applied to the CNN-derived scores. Classical ML models included *LGBM v4.1.0, SVM*, and *logistic regression* implemented using *scikit-learn v1.2.2*. Unlike the CNN, which operated directly on raw 3D MRI volumes, these models were trained on T1-derived IDPs. For each SNP, models were trained to estimate genotype class probabilities (0, 1, 2) using the same subject pool, genotype labels and train-test splits as the CNN. Predicted probabilities from the held-out test set were treated as ML-derived brain-genotype scores and analysed for their associations with cognitive phenotypes using the same linear regression framework, covariates, nested model comparisons (F-tests), and (Δ*R*^2^) measures described in Section 2.4. All models used identical data splits and matched sample sizes (Supplementary Table 1). IDPs were normalised using the UK Biobank head-size scaling factor, and class imbalance was addressed using `class weight=‘balanced’` during SVM and LGBM training. As a second comparison, cognition association analyses were repeated using the actual SNP genotype dosage (encoded as 0, 1, or 2) in place of the brain-genotype scores while controlling for the same covariates. These analyses assessed whether genotype probability scores derived from feature-based ML models, or the direct use of genotype dosage, could reproduce the associations observed with CNN-derived brain-genotype scores. To provide a degree-of-freedom-matched comparison with actual genotype dosage, we additionally performed a sensitivity analysis using the CNN-derived expected genotype dosage defined above as a single predictor. The same cognitive regression framework and covariates were applied, resulting in a 1-df test equivalent to that used for actual genotype dosage. Full details are provided in Supplementary File

### CNN model explainability

To interpret how the CNN generated brain-genotype scores, we applied the SmoothGrad method [40] with the NoiseTunnel implementation in *Captum* (v0.6.0) for PyTorch to generate voxel-wise saliency maps. Saliency maps were computed for all correctly predicted subjects in the test set and averaged to produce population-level saliency maps. To enhance interpretability, the averaged maps were thresholded to retain only the top 10% of saliency values, highlighting the brain regions contributing most strongly to the generation of brain-genotype scores. For quantitative interpretation, we used the Harvard-Oxford cortical and subcortical atlases [9], implemented in *nilearn v0.10.1* [1]. For each region of interest (ROI), we computed the mean and standard deviation of retained saliency values, together with the proportion of ROI volume covered by saliency voxels. Saliency analyses were conducted for the five SNPs whose brain-genotype scores showed the strongest associations with cognitive performance. These saliency maps were not directly associated with cognitive analyses, but were used to understand CNN decision-making and support the biological plausibility of the resulting brain-genotype scores.

## Results

### CNN Model Performance and Specification

The multi-task CNN generated brain-genotype scores for 120 SNPs analysed in groups of ten per model, using shared feature extraction layers and task-specific output heads. Scores were derived exclusively from the held-out test set and used in subsequent analyses. ResNet-10 was selected based on stable training behaviour without clear evidence of overfitting (Supplementary Figure 1-2). Alternative architectures, including deeper variants, showed less stable training dynamics and occasional indications of overfitting. Brain-genotype scores were generally well calibrated, with low top-label expected calibration error across tasks and models (median ECE = 0.016; Supplementary Figure 3), supporting their use as probabilistic biomarkers. The relationship between the probabilistic brain-genotype scores and actual genotypes was further examined using expected genotype dosage. Across pooled SNP–participant observations, CNN-derived expected dosage was positively correlated with actual genotype dosage (*r* = 0.30), with progressively higher expected scores across actual genotype classes 0, 1, and 2 (Supplementary Figure 5). This relationship indicates that the MRI-derived probability scores retain information related to the corresponding actual genotypes, while remaining continuous rather than representing hard genotype classifications. To further evaluate the generalisability of the architecture, the multi-task CNN framework was applied to sex classification jointly with nine SNP prediction tasks. The model achieved a balanced accuracy of 98.2%, a Matthews correlation coefficient (MCC) of 0.97, and an F1 score of 0.98 in the test set, indicating that the framework can support simultaneous multi-task biological prediction.

### Association of Brain-Genotype Scores with Cognitive Phenotypes

Across all brain-genotype score–cognition associations (*n* = 840), 147 remained significant after false discovery rate (FDR) correction (p*<*0.05). These involved 82 unique CNN-derived brain-genotype scores out of 120 tested, indicating that several scores were associated with multiple cognitive outcomes. Bonferroni thresholds are shown in the figures for reference. The ten most significant associations are summarised in Table 4, and full results are provided in Supplementary Results File. Figure 2(A) shows the relationship between effect sizes Δ*R*^2^ and statistical significance (*−* log_10_ *p*-values) across cognitive domains. In the sensitivity analysis including head volume as an additional covariate, associations between brain-genotype scores and cognitive measures remained largely consistent (Figure 2(B)). 50 unique SNP scores retained FDR-significant associations with at least one cognitive measure, most overlapping with the primary analysis, while five additional SNP scores became significant only after head-volume adjustment (see Supplementary Results File). However, the pattern of associations changed whereas the primary analysis identified prominent associations mainly with Fluid Intelligence and reaction time, the strongest associations after head-volume adjustment were predominantly with reaction time and Symbol Digit Matches. These findings indicate that head-volume adjustment did not broadly weaken the associations but altered their relative ranking and the specific SNP–cognition relationships identified (Figure 2).

**Table 4.** Top 10 false discovery rate (FDR)-significant associations between CNN-derived brain-genotype scores and cognitive phenotypes. P-values are from F-tests (2 df) comparing full and reduced nested models (see Methods). Full results are available in Supplementary Results File.

| SNP | Gene | Cognitive Score | p_value | $\Delta R^2$ |
| --- | --- | --- | --- | --- |
| rs13093031 | *EPHA3* | log reaction time | $9.29 \times 10^{-9}$ | 0.009 |
| rs6079584 | *MACROD2* | Symbol digit matches | $1.88 \times 10^{-8}$ | 0.010 |
| rs11699259 | *NKX2-2* | Symbol digit matches | $7.53 \times 10^{-7}$ | 0.008 |
| rs2360675 | *KLF7* | Fluid intel score | $7.75 \times 10^{-7}$ | 0.007 |
| rs763660 | *MACROD2* | Symbol digit matches | $2.40 \times 10^{-6}$ | 0.007 |
| rs3828616 | *MAP1B* | Fluid intel score | $2.93 \times 10^{-6}$ | 0.006 |
| rs2777763 | *TLE1* | Fluid intel score | $4.10 \times 10^{-6}$ | 0.006 |
| rs10846507 | *SBNO1* | log reaction time | $5.35 \times 10^{-6}$ | 0.006 |
| rs4779931 | *OTUD7A* | Fluid intel score | $7.04 \times 10^{-6}$ | 0.006 |
| rs12620095 | *TNP1* | Fluid intel score | $9.46 \times 10^{-6}$ | 0.006 |

**Figure 2.**
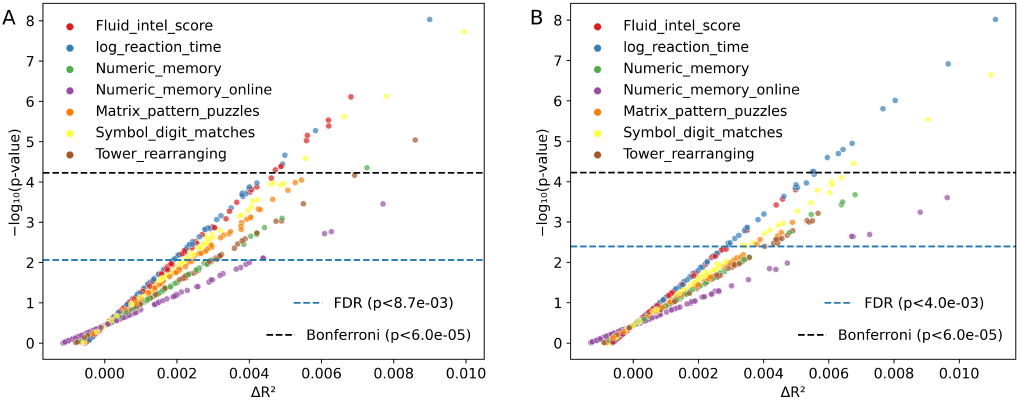
Effect size (Δ*R*^2^) versus significance (*−* log_10_ *p*) for associations of CNN-derived brain-genotype scores and cognitive phenotypes. (A) Main analysis including age, sex, education, and genetic principal components as covariates. (B) Sensitivity analysis with head-size scaling added as an additional covariate. Both panels illustrate the distribution of association strength and statistical significance across cognitive domains, showing the robustness of brain-genotype–cognition associations to head-volume adjustment. Horizontal lines indicate Bonferroni and false discovery rate (FDR) significance thresholds. **Alt text:** Scatter plots showing the relationship between effect size (Δ*R*^2^) and statistical significance (*−* log_10_ *p*) for associations between CNN-derived brain-genotype scores and cognitive phenotypes. Points represent individual SNP–cognition associations and are coloured according to cognitive test. Panel A shows the primary analysis adjusted for age, sex, education, and genetic principal components. Panel B shows a sensitivity analysis that additionally adjusts for head-size scaling. Significant associations are observed across multiple cognitive domains. Horizontal lines indicate Bonferroni and false discovery rate (FDR) significance thresholds.

### Comparative Analyses: CNN-Derived Brain-Genotype Scores vs Classical Models and Actual Genotypes

CNN-derived brain-genotype scores outperformed both classical machine learning models and actual genotype data in explaining cognitive phenotypes. Classical models trained on IDPs (SVM, LGBM, and logistic regression) yielded few significant associations after multiple-testing correction. Only one LGBM-derived score (rs61966099) was significantly associated with fluid intelligence (*p* = 5.24 *×* 10^*−*5^) after FDR correction (Figure 3). Similarly, when actual genotype data were analysed directly, only one SNP (rs2360675) showed a significant association with reaction time (*p* = 4.15 *×* 10^*−*5^); Supplementary Results File. Supplementary Figures 7–8 further illustrate these comparisons through direct contrasts of association significance and effect sizes obtained using CNN-derived brain-genotype scores and actual genotypes. To provide a degree-of-freedom-matched comparison with actual genotype dosage model, we additionally tested CNN-derived expected genotype dosage as a single predictor (1 df). The expected-dosage analysis retained stronger cognition associations than actual genotype dosage and showed broadly comparable overall effect sizes to the primary probability-based analysis, although individual association patterns differed (Supplementary Figure 6).

**Figure 3.**
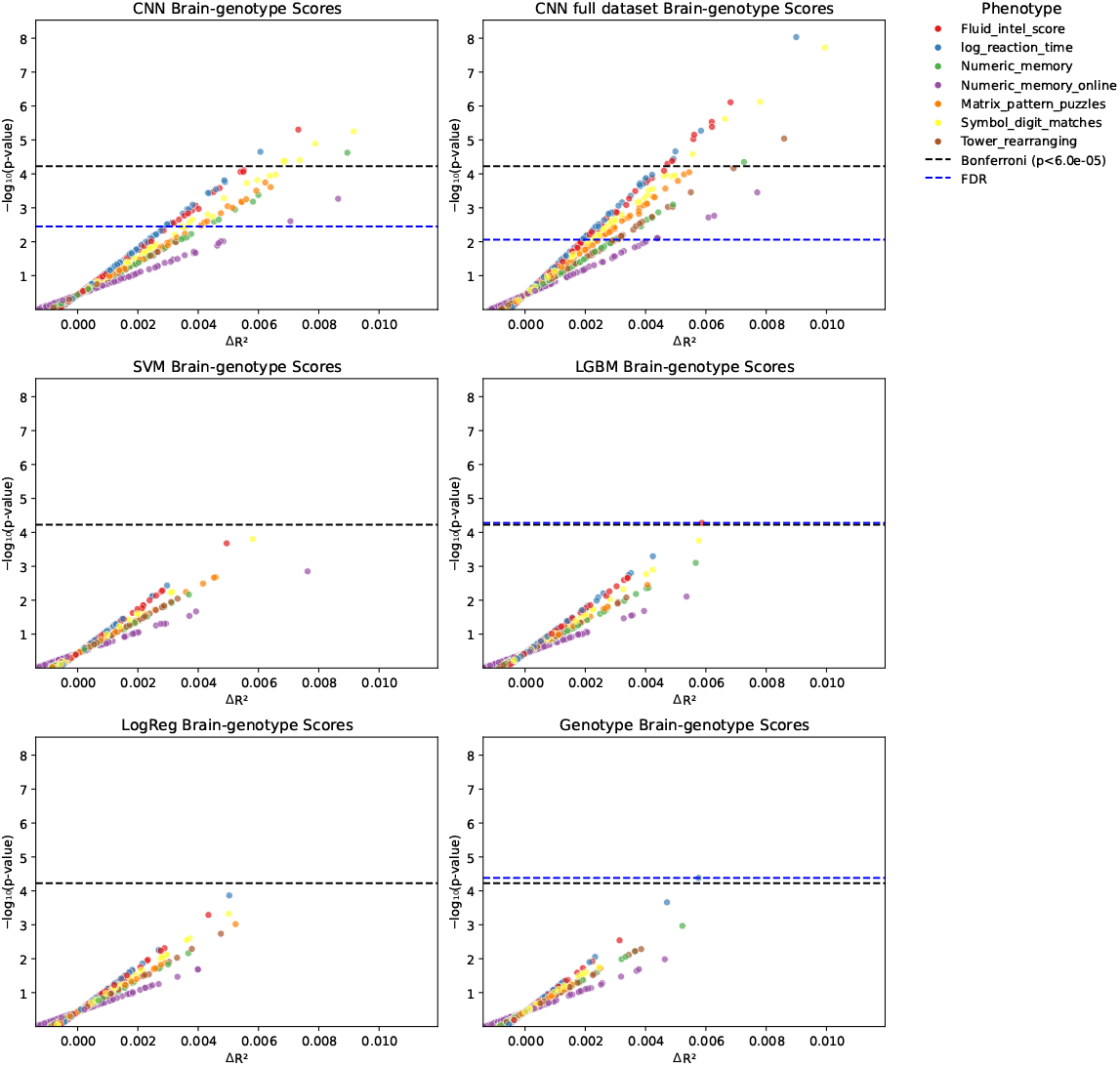
Effect size (Δ*R*^2^) versus significance (*−* log_10_ *p*) for associations between brain-genotype scores and cognitive phenotypes across modelling approaches. Each panel corresponds to a different model: CNN, SVM, LGBM, logistic regression, and actual genotype data. Two CNN panels are included: the first uses the identical dataset as the alternative models (SVM, LGBM, logistic regression, and actual genotype data), ensuring direct comparability. The second CNN panel (CNN full dataset) leverages a larger dataset, highlighting the improved strength and significance of genotype-cognition associations achievable with increased sample size. **Alt text:** Multi-panel scatter plots comparing associations between brain-genotype scores and cognitive phenotypes across modelling approaches. CNN-derived scores show more significant associations and larger effect sizes than support vector machine, Light Gradient Boosting Machine, logistic regression, or actual genotype data, with the strongest associations observed for the CNN model trained on the larger dataset.

### Reproducibility of SNP scores

Cross-seed reproducibility of CNN-derived scores showed substantial heterogeneity across SNPs. While many SNPs exhibited high correlations across confidence thresholds, others showed moderate, low, or occasionally negative correlations, indicating variable stability of brain-genotype scores across model initialisations (Supplementary Figure 9). These findings suggest that the CNN captures robust brain-derived signals for a subset of SNPs, whereas other scores may reflect weaker or less stable neuroimaging signals. Mean correlation values did not differ significantly between SNPs that were significantly associated with cognitive phenotypes and those that were not (Mann–Whitney U = 1626, p = 0.70). However, a permutation test suggested a trend towards greater reproducibility among significant SNPs (p = 0.054), although this did not reach conventional significance thresholds (Figure 4).

**Figure 4.**
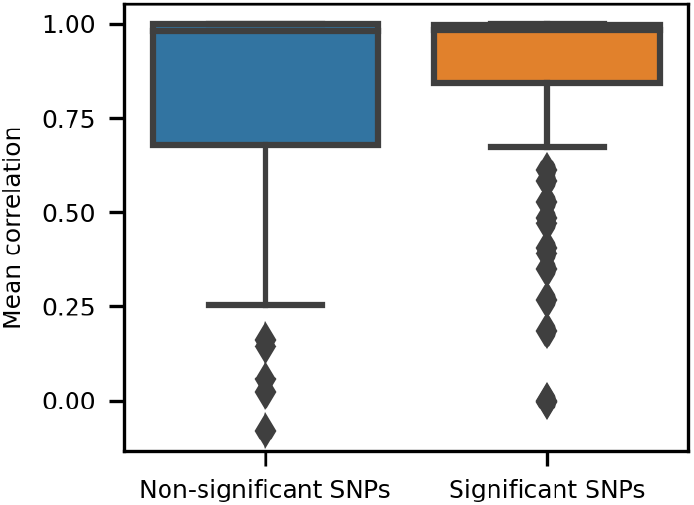
Distribution of mean seed-to-seed correlations for CNN-derived SNP probability biomarkers. Boxplots show reproducibility of the SNP-level probability vectors between two model seeds for SNPs that were significantly (FDR-corrected p*<*0.05) or non-significantly associated with cognitive phenotypes. **Alt text:** Boxplots comparing the reproducibility of CNN-derived brain-genotype scores between significant and non-significant SNPs. Both groups show generally high correlations with overlapping distributions.

### CNN Model Explainability

Population-level saliency maps identified the brain regions contributing most strongly to the generation of CNN-derived brain-genotype scores. Representative saliency maps for the top SNPs showing the strongest associations with cognitive measures are shown in Figure 5, and quantitative summaries across Harvard–Oxford cortical and subcortical regions are presented in Figure 6. Across SNPs, saliency maps displayed distinct yet partially overlapping spatial patterns, indicating that the multi-task CNN learned SNP-specific neuroanatomical representations while capturing shared biological substrates. Specifically, ventricular regions (left and right lateral ventricles) consistently ranked among the most salient regions across all SNPs. In addition, subcortical regions including the hippocampus, thalamus, and caudate nucleus were repeatedly identified among the top-ranked regions across multiple SNPs, indicating overlap in regions contributing to CNN decision-making. Cortical regions and cerebral white matter also showed recurrent saliency coverage across SNPs, although with lower relative rankings compared to ventricular and subcortical structures.

**Figure 5.**
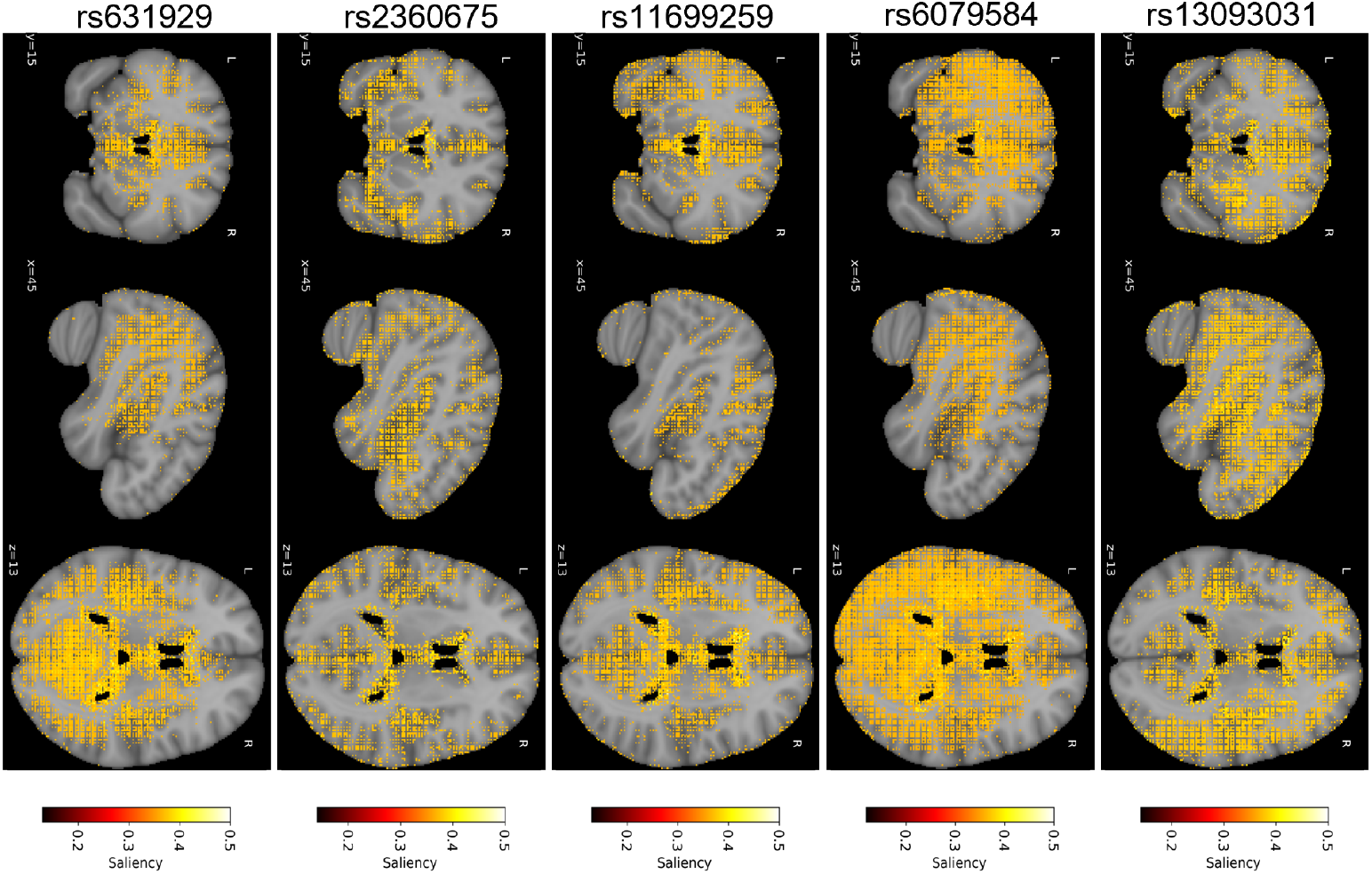
Average saliency maps for the top brain-genotype scores across three orthogonal slices: axial, coronal, and sagittal. **Alt text:** Brain images showing average saliency maps for the five brain-genotype scores with the strongest associations to cognitive phenotypes. Axial, coronal, and sagittal views are shown for each score. Highlighted regions indicate brain areas that contributed most strongly to CNN genotype prediction, with distinct but partially overlapping spatial patterns across SNPs.

**Figure 6.**
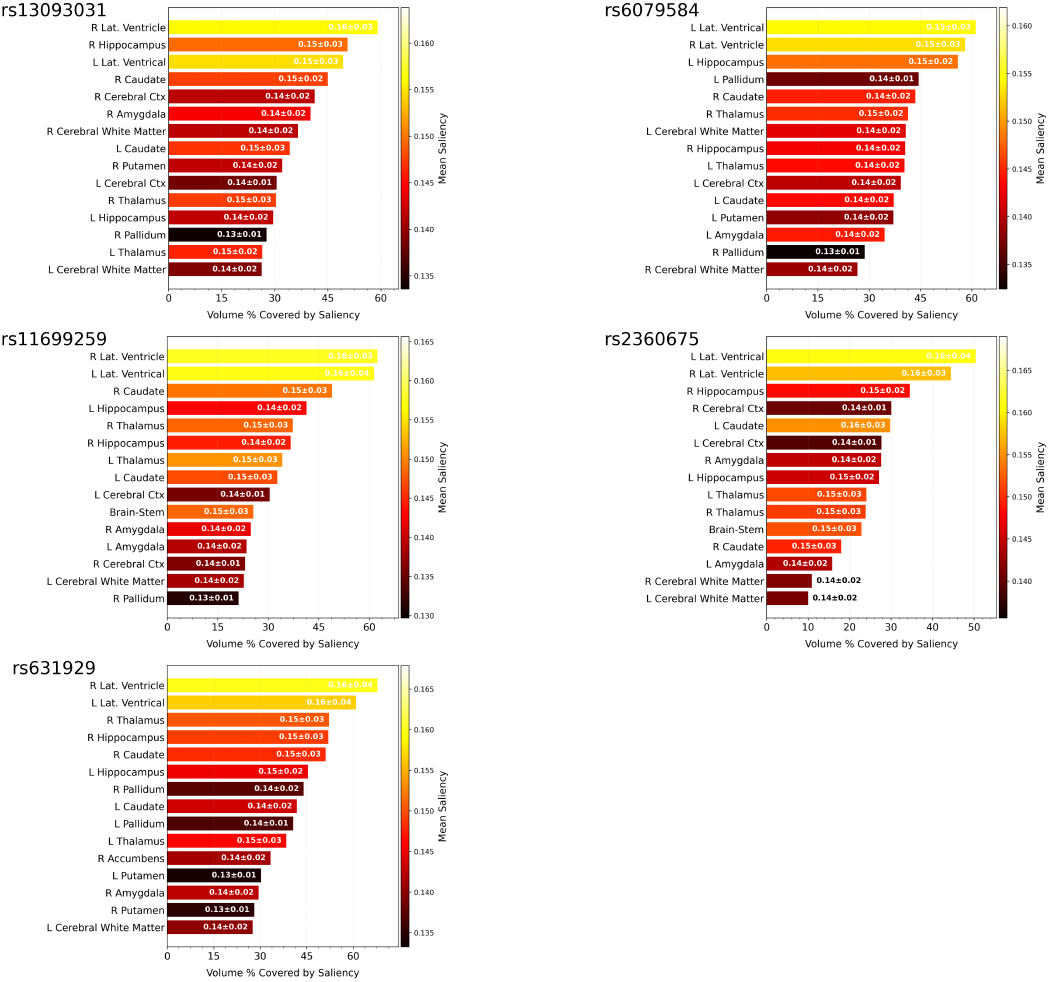
Saliency mean values and standard deviations for cortical and subcortical brain regions associated with top brain-genotype scores. **Alt text:** Bar charts showing mean saliency values for cortical and subcortical brain regions associated with the five most significant brain-genotype scores. Ventricular regions, hippocampus, thalamus, caudate, and other subcortical structures are among the most frequently highlighted regions contributing to CNN genotype prediction.

## Discussion

Traditional imaging genetics approaches, which rely on predefined imaging-derived phenotypes and linear statistical models, have advanced the field but remain limited in capturing the complex ways genetic variation shapes brain structure. By reducing high-dimensional MRI data into summary features, such methods can overlook distributed and non-linear genotype effects expressed across interconnected brain regions. To address these limitations, we developed an explainable multi-task convolutional neural network (CNN) framework that learns SNP-specific brain-genotype relationships directly from raw MRI data. The resulting brain-genotype scores provide probabilistic neuroanatomical representations of genetic variation, offering a data-driven link between genotype and brain anatomy. Conceptually, this framework shifts imaging genetics from localised, predefined features towards distributed representations of genetic effects, consistent with the polygenic and network-based organisation of the brain.

Across cognitive phenotypes, CNN-derived brain-genotype scores showed substantially more and stronger associations than scores derived from classical machine learning approaches (147 associations compared to 1), while associations using actual genotype data were largely absent. This finding suggests that brain-genotype scores capture brain-mediated components of genetic variation that are more closely related to cognition than genotype alone. Importantly, these associations emerged without explicitly training the CNN on cognition outcomes, indicating that the learned representations reflect biologically meaningful structure rather than task-specific overfitting. As such, the framework may serve as a discovery-oriented tool for identifying genetic variants whose effects on cognition are mediated through distributed brain anatomy.

A key aspect of this approach is the use of probabilistic outputs rather than hard genotype classifications. Probability-based brain-genotype scores reflect the subtle, continuous nature of genotype-related neuroanatomical variation, particularly in a healthy, middle-aged population where individual genetic effects are small and spatially diffuse. Prior reverse imaging–genetics studies have often focused on variants with larger effects, such as APOE [6, 11, 27], often in case–control cohorts [34, 24, 19, 46], where disease-related changes amplify signal differences. In contrast, our approach models normative variation, enabling the detection of graded genetic influences on brain structure that are not well captured by categorical or linear models. The multi-task design further strengthens this framework by allowing the CNN to learn shared representations across multiple SNPs while preserving SNP-specific spatial signatures. By sharing information across tasks, the model reduces sensitivity to SNP-specific noise and provides a biologically grounded framework for identifying distributed genotype–brain relationships beyond the capacity of univariate or linear models. Our association analyses identified several SNPs whose brain-genotype scores were significantly linked to multiple cognitive measures. One of the top findings, rs13093031, mapped to the gene *EPHA3*, was associated with reaction time, and symbol digit matching, both of which primarily measure processing speed, as well as matrix pattern puzzles, which reflect fluid intelligence [37]. While rs13093031 itself has not previously been directly associated with cognition, the *EPHA3* gene has been reported in the literature to be associated with intelligence, using several datasets including UK Biobank cognitive phenotypes such as fluid intelligence [36], regional brain volumes [47], and functional connectivity networks through associations with rs-fMRI measures [49], thereby reinforcing the biological plausibility of this result. Similarly, rs2360675 in the *KLF7* gene showed associations with all cognitive phenotypes and was the only SNP whose actual genotype data showed a significant, though weaker, relationship with cognition. This SNP has previously been reported in the literature as associated with brain volume [47], poorer self-rated health [28, 14], and age-related hearing loss [33].

Beyond SNP-specific findings, the observed cognitive patterns should be interpreted in the context of the brain-genotype scores, which encode genetic variation as it is expressed in distributed brain structure. In the main analysis, fluid intelligence and reaction time consistently showed larger effect sizes and more frequent significant associations than other cognitive measures. In sensitivity analyses adjusting for head volume, associations with fluid intelligence were substantially attenuated, whereas reaction time and symbol digit matching—both measures of processing speed—emerged as the most robust and consistently associated outcomes.

This shift suggests that some genotype–cognition relationships observed in the main model may be mediated through global brain size, consistent with evidence linking intelligence to overall brain volume [32]. In contrast, associations with processing speed appear to reflect distributed neuroanatomical patterns that remain after accounting for head volume, in line with prior work showing strong links between processing speed and brain-wide white matter integrity and neural efficiency [31].

To characterise the neuroanatomical substrates underlying these associations, saliency maps were used to identify brain regions contributing most strongly to SNP-specific score generation. Importantly, these maps were derived independently of any cognition association analyses and therefore reflect the anatomical features used by the CNN to estimate brain-genotype scores, rather than post hoc explanations of cognitive effects. Although all SNPs shared a common feature extraction backbone within the multi-task CNN, the resulting saliency maps showed distinct spatial patterns across SNPs, indicating that the model learned SNP-specific neuroanatomical representations. At the same time, they still highlight partially overlapping regions that may reflect shared biological pathways.

Among SNPs whose scores were significantly associated with cognitive phenotypes, saliency maps consistently highlighted a set of core cortical and subcortical structures across variants, with SNP-specific differences in their relative contributions. The most frequent regions included the bilateral ventricles, hippocampus, caudate, cerebral cortex, amygdala, and thalamus. Many of these regions are well established in the literature as central to cognitive function [12, 21, 23]. Importantly, while the precise ordering of regional saliency varied across SNPs, the overlap of these core brain regions suggests that the CNN captures distributed neuroanatomical signatures through which genetic variation may influence cognitive performance rather than relying on isolated or region-specific effects.

### Limitations and Future Directions

Future work could extend this framework in several ways. Longitudinal data could help assess temporal dynamics and causal pathways linking genetic variation, brain structure, and cognition. Applying the framework to multimodal data (e.g., diffusion MRI, fMRI, or blood-based biomarkers) may improve the sensitivity of brain-genotype scores by integrating complementary sources of biological information. Replication in independent cohorts is also needed to confirm the robustness of the identified SNP–cognition associations. As a proof-of-concept, this study focused on 120 SNPs previously identified in imaging GWAS as associated with T1-derived phenotypes, to evaluate the feasibility of deriving brain-genotype scores for variants with known imaging relevance. Extending the framework to genome-wide analysis represents a natural next step, where CNN-derived scores could be generated for all SNPs and tested for associations with cognitive or clinical phenotypes in a reverse GWAS design. Such large-scale implementation would enable systematic discovery of genotype–brain–behaviour relationships on a genome-wide scale. By capturing distributed neuroanatomical representations of genetic variation, brain-genotype scores may serve as intermediate phenotypes for future studies linking genetic architecture to cognitive trajectories or disease risk.

## Conclusions

In this study, we introduced brain-genotype scores, image-based representations of genetic variation learned directly from structural MRI. These scores showed stronger associations with cognitive phenotypes than actual genotypes or IDP-based machine learning approaches, demonstrating their utility for identifying brain-mediated genetic effects.

## Data Availability

All data produced in the present study are available upon reasonable request to the authors

## Author contributions

KTA conceptualised the study, developed the analysis code, performed the analyses, and wrote the manuscript. ANH provided primary supervision and contributed to study design and interpretation of results LW, BM, CVD provided ongoing supervision, contributed to the interpretation of the findings The remaining authors contributed to technical support, computational infrastructure, and manuscript revision. All authors have read and agreed to the published version of the manuscript.

## Data availability

UK Biobank data is available via application on the cohort website.

https://www.ukbiobank.ac.uk/enable-your-research/apply-for-access

## Acknowledgments

This work was supported by the Centre of Artificial Intelligence in Precision Medicines (CAIPM), King Abdulaziz University, Jeddah, Saudi Arabia. This research has been conducted using the UK Biobank Resource under Application Number 15181.

## Biographical Note

Kholod Thaker Alhasani is a doctoral researcher at the University of Oxford, working on imaging genetics, neuroimaging, and artificial intelligence methods for precision medicine.

