## Supplementary_Material for "An Explainable Deep Learning Framework for Imaging Genetics: Deriving Brain-Genotype Scores to Link Genetic Variation, Brain Structure, and Cognition"

### Model Validation and Calibration of CNN-Derived Brain–Genotype Scores

The multitask ResNet10 models and their output probability scores were evaluated with respect to training stability, probabilistic calibration, and discriminative performance. As the outputs are used as probabilistic biomarkers, evaluation focused on the properties of the learned brain–genotype scores across SNPs rather than single-task classification accuracy, given the expected small effect sizes of individual SNPs on brain anatomy.

#### Training Stability

Training stability was assessed using training and validation cross-entropy loss with early stopping based on validation loss. At each epoch, the total loss was computed as the sum of cross-entropy losses across the 10 SNP prediction tasks and used to update the model parameters. Across all 12 models, both training and validation losses showed an initial decrease followed by early stabilisation, with no evidence of divergence between training and validation curves (Figure 1). Convergence was generally modest, consistent with the low signal-to-noise ratio expected in SNP–brain associations. Representative task-level loss curves are shown in Figure 2. These illustrate stable optimisation dynamics with moderate stochastic fluctuations, reflecting weak genetic signal without evidence of model overfitting.

#### Calibration of Brain–Genotype Scores

Calibration was assessed using reliability diagrams and expected calibration error (ECE) [1]. The aim was to evaluate whether predicted probabilities align with observed genotype frequencies. ECE was computed per SNP and summarised across predicted SNPs within each model. Results are shown as box plots in Figure 3, where each box represents the distribution of ECE across the 10 tasks in a model. ECE ranges from 0 (perfect calibration) to 1 (poor calibration). The median ECE across tasks was 0.016. Representative reliability diagrams are shown in Figure 4. These were computed by aggregating predictions across tasks within each model to obtain stable estimates. Curves are shown for each genotype and the top predicted class. Alignment with the diagonal indicates good calibration; deviations below the diagonal indicate overconfidence, and deviations above indicate underconfidence. Overall, the predicted genotype probabilities demonstrated reasonable agreement with observed frequencies, indicating well-calibrated probabilistic scores suitable for use as biomarkers.

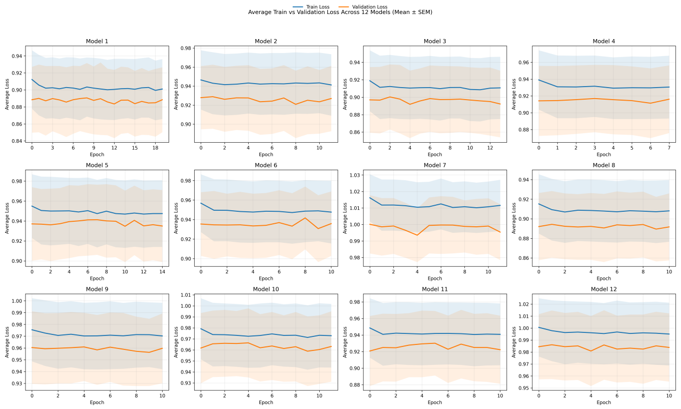


**Figure 1** Training and validation loss across models. Each panel shows training and validation loss per epoch for one model. For visualisation, loss is reported as the average across the 10 SNP tasks within each model, while model optimisation was performed using the sum of task losses.

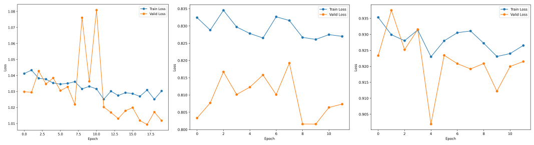

**Figure 2** Three representative task-level training and validation loss curves. Training and validation loss per epoch for selected SNP genotype prediction tasks. These examples illustrate typical task-level behaviour, including stable convergence with moderate stochastic fluctuations due to low signal.

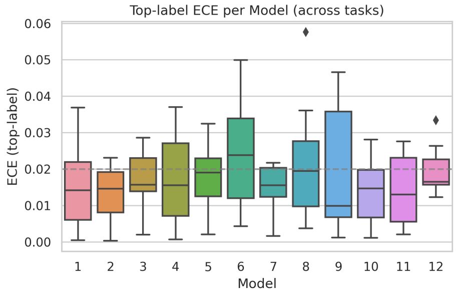


**Figure 3** Top-label Expected Calibration Error (ECE) across tasks for each model. Each box summarises the distribution of ECE values over tasks, with lower values indicating better calibration. Most models exhibit low calibration error (median ECE  0.016), suggesting generally well-calibrated predictions, while variability within boxes reflects task-dependent differences in calibration performance.

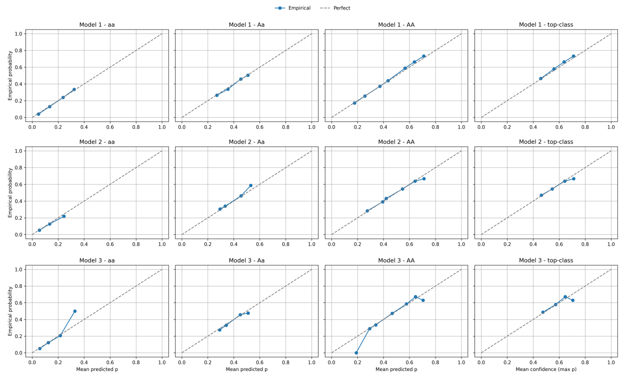

**Figure 4** Representative calibration curves across models. Reliability diagrams for three representative models, computed by aggregating predictions across all SNP tasks within each model. Curves are shown for each genotype (aa, Aa, AA) and the top predicted class. The dashed line indicates perfect calibration. Overall, predicted probabilities are reasonably aligned with observed frequencies.

### Relationship Between CNN-Derived Expected Genotype Dosage and Observed Genotype Dosage

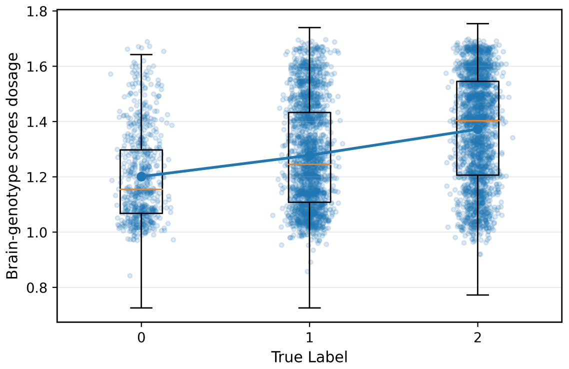


**Figure 5** Relationship Between CNN-Derived Expected Genotype Dosage and Observed Genotype Dosage. Brain–genotype scores were summarised as expected genotype dosage, calculated from the three CNN-generated genotype probabilities $0 \times p_0 + 1 \times p_1 + 2 \times p_2$. Distributions are shown for participants grouped by observed genotype class (0, 1, or 2). Points represent individual participants, boxplots show within-class distributions, and connected points indicate class-specific means. Across all SNP scores and observed genotypes, expected dosage showed a positive Pearson correlation ($r = 0.30$). The progressive increase in expected dosage across observed genotype classes further demonstrates that the MRI-derived scores retain information related to the corresponding observed genotypes.

### Sensitivity Analysis Using CNN-Derived Expected Genotype Dosage

To assess the robustness of the brain-genotype score associations to an alternative representation of the CNN outputs, the CNN-derived expected genotype dosage defined above was used as a single continuous predictor in the cognition association analyses. The same participants, cognitive phenotypes, covariates, and nested regression framework as in the primary analysis were retained. Unlike the primary analysis, which jointly included two independent probability components using a 2-df F-test, the expected-dosage analysis used a 1-df test. This formulation also enabled a degree-of-freedom-matched comparison with the observed genotype dosage analysis. The CNN-derived expected-dosage analysis retained stronger cognition associations than the observed genotype dosage analysis, while showing broadly comparable overall effect sizes to the primary two-component probability analysis. However, the pattern of individual SNP–cognition associations was not identical to the primary analysis, with some cognitive outcomes (Reaction Time) showing greater consistency across score representations than others. These findings indicate that the overall association signal was retained when the CNN probabilities were reduced to a single expected-dosage predictor, while some information captured by the full probability representation was not preserved by this one-dimensional summary

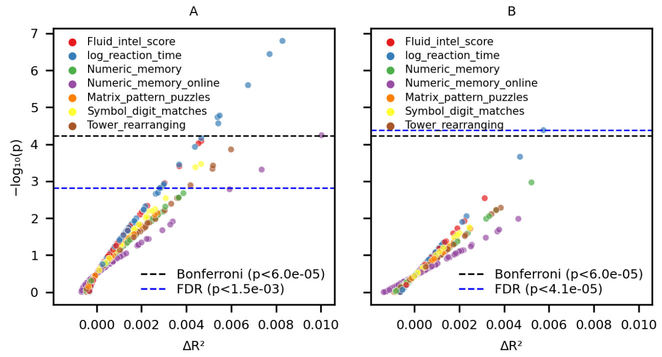


**Figure 6** Comparison of cognition associations using CNN-derived expected genotype dosage and observed genotype dosage. (A) Associations between CNN-derived expected genotype dosage and seven cognitive phenotypes. Expected dosage was derived from the CNN-generated genotype probabilities and entered as a single continuous predictor. (B) Corresponding associations using observed genotype dosage (0, 1, or 2) as a single predictor. Both analyses used the same regression framework and covariates, providing a degree-of-freedom-matched (1-df) comparison. Each point represents an individual SNP–cognition association, with colours indicating cognitive phenotype. The x-axis shows the additional variance explained by the dosage predictor ($\Delta R^2$), and the y-axis shows statistical significance ($-\log_{10}(p)$). Horizontal dashed lines indicate the false discovery rate (FDR) and Bonferroni significance thresholds for each analysis.

### CNN-Derived Brain–Genotype Scores vs Classical Models and Actual Genotypes

**Table 1** Demographic data for the linear regression models dataset used in cognitive outcome analyses. This dataset corresponds to the testing set of the CNN models, SVM, LGBM, and logistic regression used to generate brain–genotype scores.

| Total $N = 3\,057$ | | |
|---|---|---|
| **Age** | | |
| mean | 64.6 | |
| std | 7.6 | |
| min | 46 | |
| 25% | 59 | |
| 50% | 65 | |
| 75% | 70 | |
| max | 82 | |
| **Sex** | **Count** | **Percentage (%)** |
| Female | 1 577 | 51.6 |
| Male | 1 480 | 48.4 |
| **Assessment Centre** | **Count** | **Percentage (%)** |
| 11025 | 1 780 | 58.2 |
| 11027 | 807 | 26.4 |
| 11026 | 465 | 15.2 |
| 11028 | 5 | 0.2 |

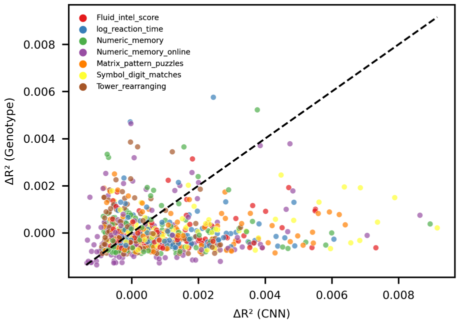


**Figure 7** Scatter plot compares $\Delta R^2$ from from linear models using CNN-derived brain–genotype scores vs. actual genotypes in association with cognitive phenotypes.

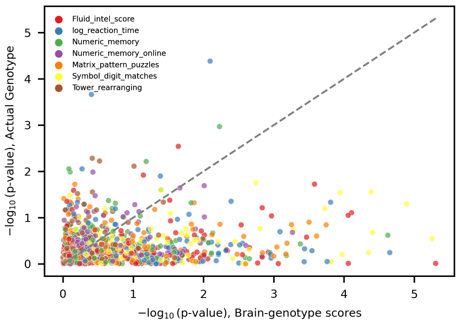


**Figure 8** Scatter plot compares the $-\log_{10} p$ from linear models using CNN-derived brain–genotype scores vs. actual genotypes in association with cognitive phenotypes.

### Reproducibility of brain–genotype scores

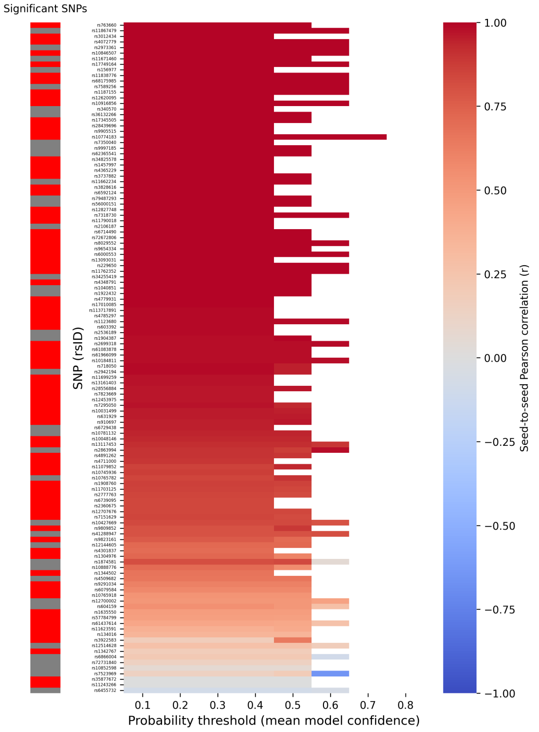


**Figure 9** Reproducibility of CNN-derived SNP probability biomarkers across random seeds. The heatmap (right) shows Pearson correlation coefficients between two model seeds for the three-class probability outputs of each SNP, calculated at increasing probability thresholds (x-axis). Higher thresholds correspond to including only subjects with higher average model confidence. Each row corresponds to one SNP (rsID), and the colour scale represents the strength of seed-to-seed correlation (r). The side bar (left) indicates SNPs that were significantly associated with cognitive phenotypes (p¡0.05, red = significant, gray = non-significant).
